# Predict multicategory causes of death in lung cancer patients using clinicopathologic factors

**DOI:** 10.1101/2020.09.25.20201095

**Authors:** Fei Deng, Haijun Zhou, Yong Lin, John Heim, Lanlan Shen, Yuan Li, Lanjing Zhang

## Abstract

**Background:** Random forest model is a recently developed machine-learning algorithm, and superior to other machine learning and regression models for its classification function and better accuracy. But it is rarely used for predicting causes of death in lung cancer patients. On the other hand, specific causes of death in lung cancer patients are poorly classified or predicted, largely due to its categorical nature (versus binary death/survival).

**Methods:** We therefore tuned and employed a random forest algorithm (Stata, version 15) to classify and predict specific causes of death in lung cancer patients, using the surveillance, epidemiology and end results-18 and several clinicopathological factors. The lung cancer diagnosed during 2004 were included for the completeness in their follow-up and death causes. The patients were randomly divided into training and validation sets (1:1 match). We also compared the accuracies of the final random forest and multinomial regression models.

**Results:** We identified and randomly selected 40,000 lung cancers for the analyses, including 20,000 cases for either set. The causes of death were, in descending ranking order, were lung cancer (72.45 %), other causes or alive (14.38%), non-lung cancer (6.87%), cardiovascular disease (5.35%), and infection (0.95%). We found more 250 iterations and the 10 variables produced the best prediction, whose best accuracy was 69.8% (error-rate 30.2%). The final random forest model with 300 iterations and 10 variables reached an accuracy higher than that of multinomial regression model (69.8% vs 64.6%). The top-10 most important factors in the random-forest model were sex, chemotherapy status, age (65+ vs <65 years), radiotherapy status, nodal status, T category, histology type and laterality, which were also independently associated with 5-category causes of death.

**Conclusion:** We optimized a random forest model of machine learning to predict the specific cause of death in lung cancer patients using a set of clinicopathologic factors. The model also appears more accurate than multinomial regression model.

## Introduction

Lung cancer is one of the leading causes of death in the U.S. and the world [1]. Basic research has revealed new insights into development and progression of lung cancer [2]. Clinical works also identified the factors associated with the survival outcomes of lung cancer, particularly non-small cell lung cancer, including histology and oncogenic drivers [3]. However, the survival outcomes of the lung cancers are mostly binary, which were either alive versus death or alive versus progression/death. Few studies on lung cancer patients were focused on the specific deaths due to non-lung cancer[4], cardiovascular diseases[5, 6], infection or other causes[4, 7]. Even fewer works investigated the multicategory causes of death (COD).

Random forest (RF) model is a recently developed machine-learning algorithm, and superior to other machine learning and regression models for its classification function and better accuracy [8]. But it is rarely used for predicting causes of death in lung cancer patients, while we have shown a better prediction accuracy in prostate cancers [8]. On the other hand, specific causes of death in lung cancer patients are poorly classified or predicted, largely due to its categorical nature (versus binary death/survival). Therefore, this study was aimed to understand the factors associated with multicategory COD in lung cancer patients using RF and multinomial regression models.

## Material and methods

We extracted clinical, pathologic and socioeconomic data of the patients in the Surveillance, Epidemiology, and End Results-18 (SEER-18) Program (www.seer.cancer.gov) SEER*Stat Database with Treatment Data, who were diagnosed of lung carcinoma in 2004. The follow up was last conducted in Nov. 2016 (the last in the 2019 data release). The inclusion criteria included the survival time > 1 month, aged 20+ years, first primary-cases only and having a known COD. The average follow-up time was 12.5 years.

The informed consent was not obtained for the SEER patients due to de-identified nature of the dataset. Owing to the use of publicly available, de-identified SEER cases, this study was exempt from an institutional review board approval. However, we have received the approval for using the SEER-18 data under the condition of compliance with their preset terms (user ID lzhang). Moreover, all 50 states in the USA have laws requiring newly diagnosed cancers to be reported to a central registry. The state cancer registries in the SEER program would deposit their extracted, de-identified cancer data to the SEER database after meeting quality control standards (www.seer.cancer.gov). Thus, the SEER data collection was authorized by the US state laws, and supervised by respective state public-health officials and ethical review committees.

We simplified the SEER COD, which were extracted from death certificates, into 5 categories, namely cardiovascular system disease (CVS), infection, non-breast cancer, breast cancer and others cause (including alive). The alive patients were a small proportion of the population and were thus included in the other causes. We included 40 dichotomized variables in the analyses since dichotomized variables (i.e., one-hot encoding) produced slightly better accuracy as shown before[8] and would not require normalization.

We tuned and employed a random forest algorithm (Stata, version 15) to classify and predict specific causes of death in lung cancer patients. The lung cancer diagnosed during 2004 were included for the completeness in their follow-up and death causes. The patients were randomly divided into training and validation sets (1:1 match). We also compared the accuracies of the final random forest and multinomial regression models using chi-squared test. A p value <0.05 was considered statistically different.

## Results

Among the 44,735 case diagnosed in 2004, we identified and randomly selected 40,000 qualified lung cancers for the analyses, including 20,000 cases for either train or test set (**Table 1**). The causes of death were, in descending ranking order, were lung cancer (72.45%), other causes or alive (14.38%), non-lung cancer (6.87%), cardiovascular disease (5.35%), and infection (0.95%). The mean follow-up time was 114.8 months (standard deviation 49.1).

**Table 1.**
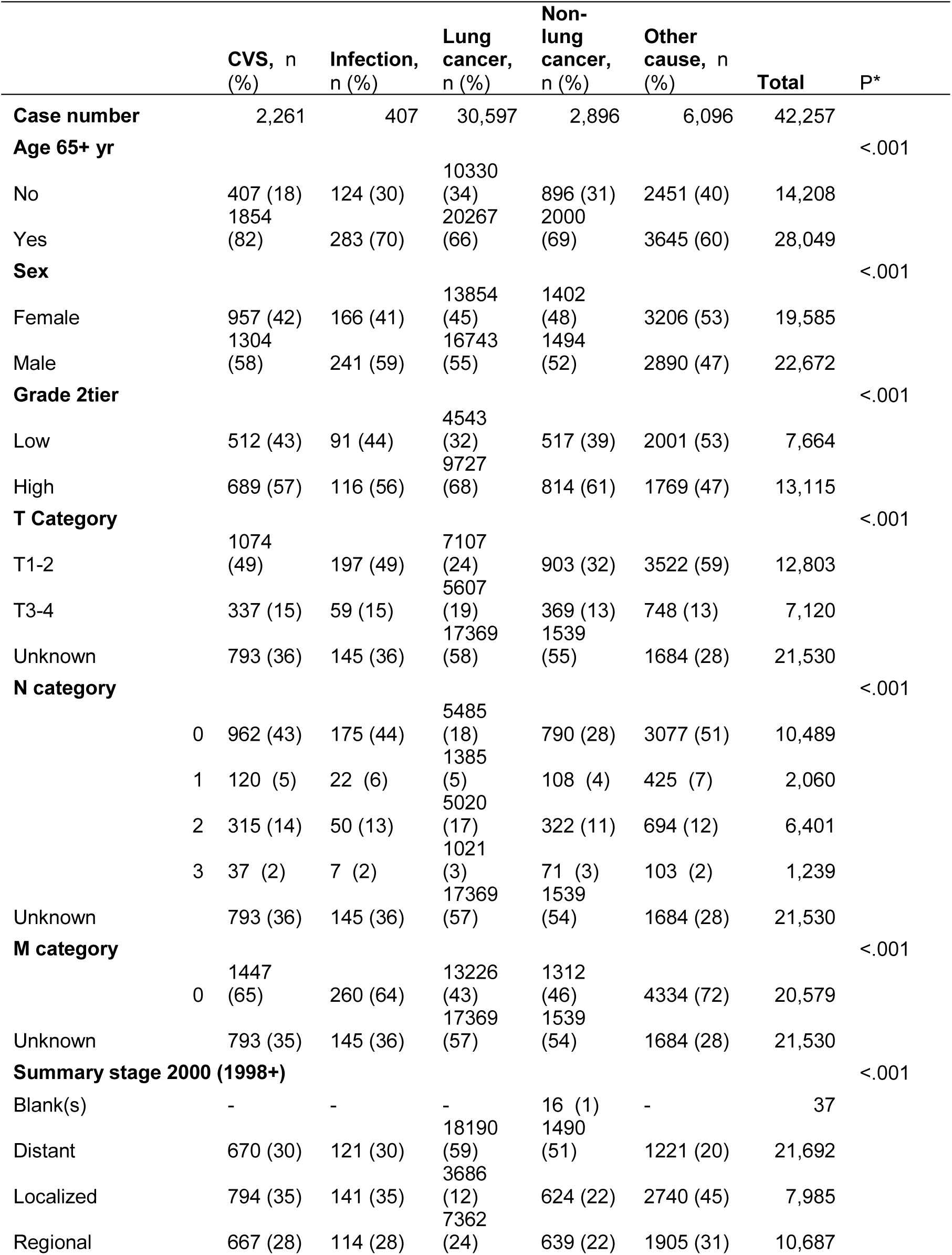
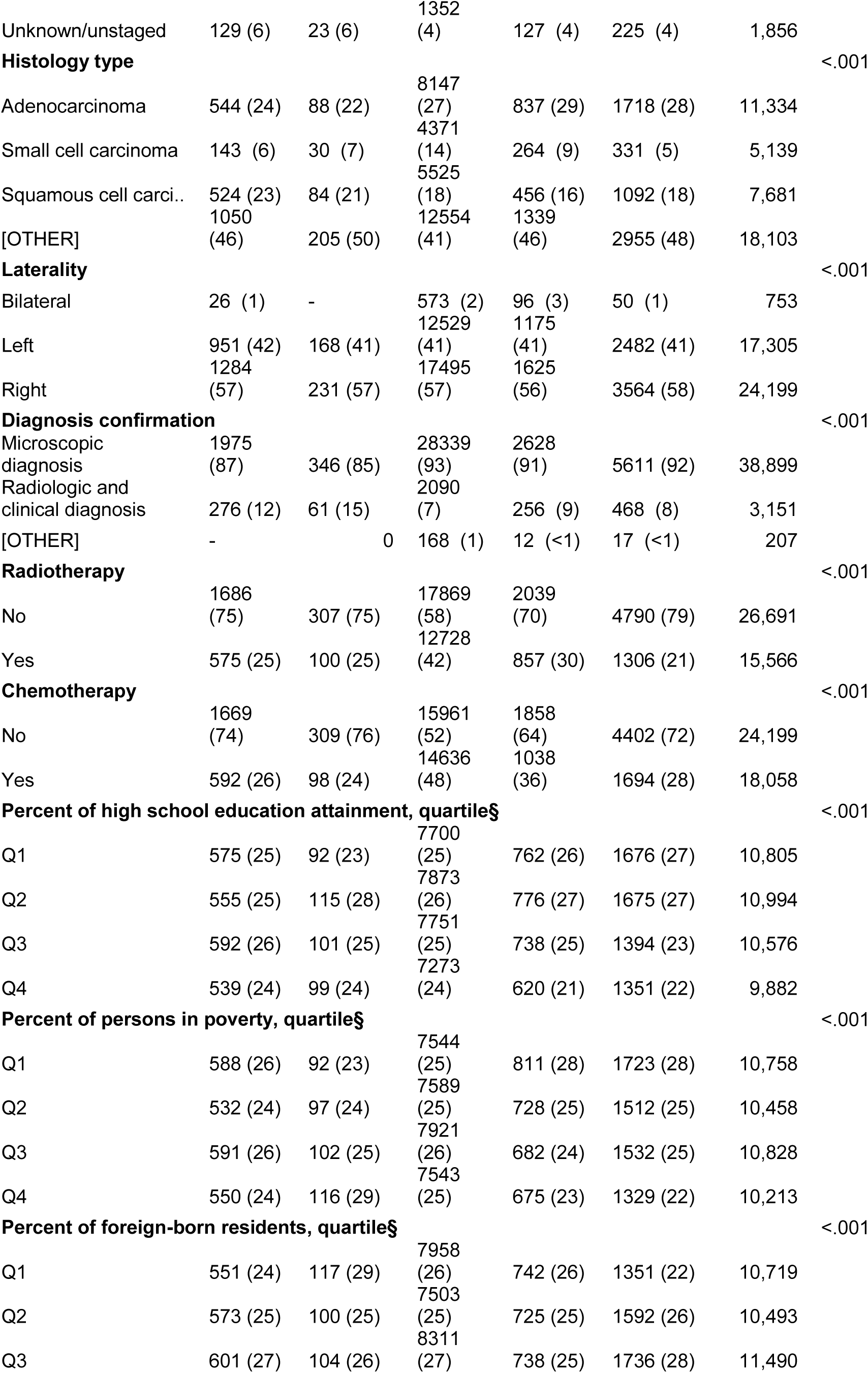
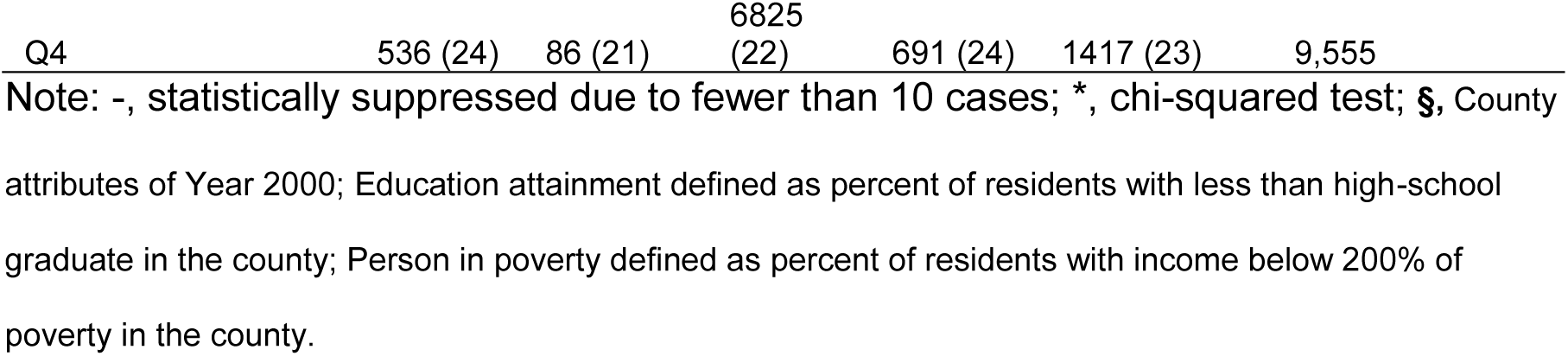
Baseline characteristics of the included cases according to the 5-category Cause of death.

We found more 250 iterations and the 10 variables produced the best prediction, whose best accuracy was 69.8% (error-rate 30.2%, **Figure 1**).

**Figure 1.**
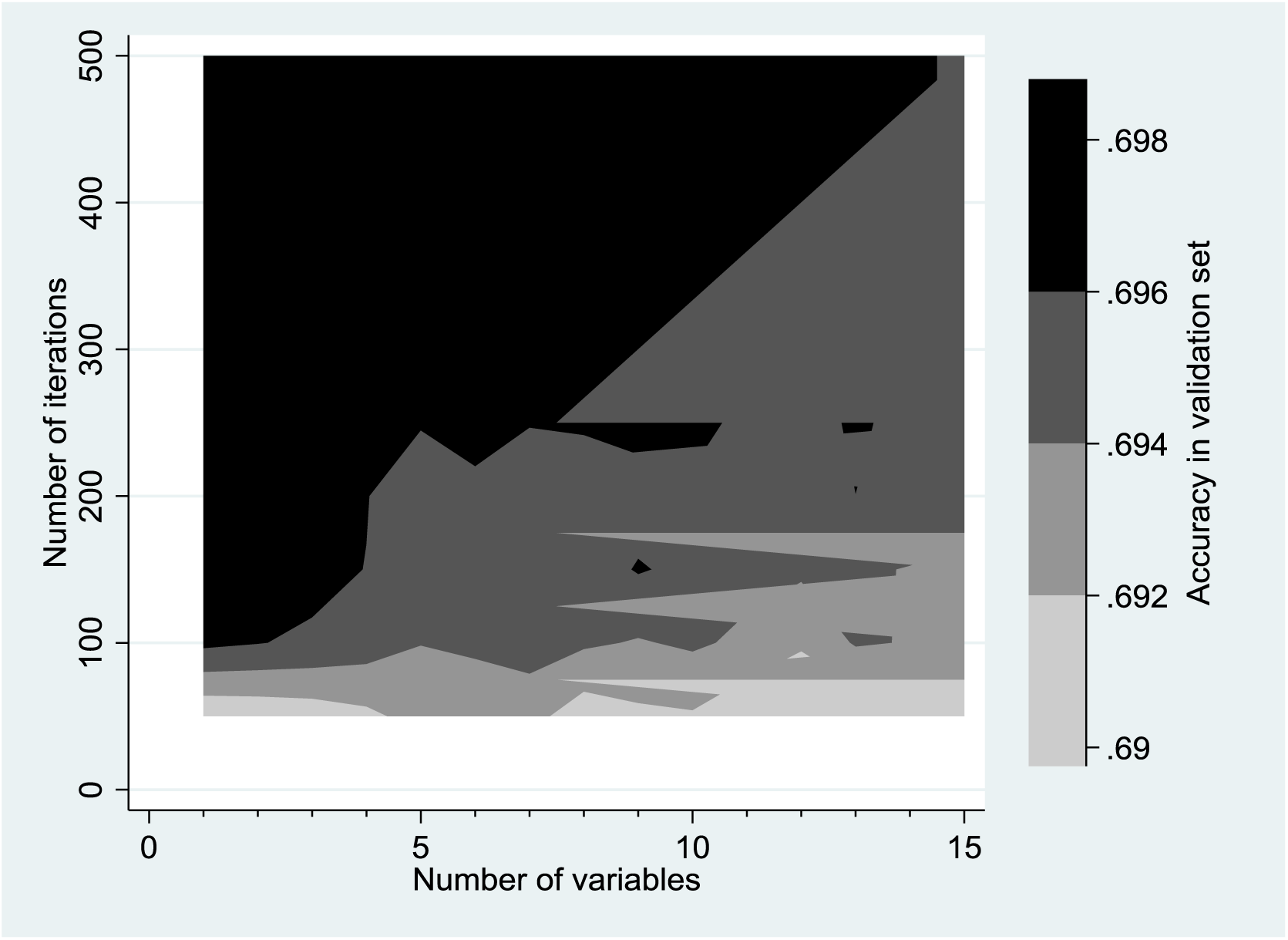
The error rates in the validation set were reduced as the number of iterations and variables increase. But more than 10 variables were linked to a higher error rate.

The final random forest model with 300 iterations and 10 variables reached an accuracy higher than that of multinomial regression model (69.8% vs 64.6%, P<0.001, **Table 2**). The top-10 most important factors in the random-forest model were sex, chemotherapy status, age (65+ vs <65 years), radiotherapy status, nodal status, T category, histology type and laterality (**Figure 2**).

**Table 2.** Confusion matrices of the random forest and multinomial regression models.

| Predicted classes | CVS | Infection | Lung cancer | Non-lung cancer | Other cause | Total |
| --- | --- | --- | --- | --- | --- | --- |
| Random forest model |  |  |  |  |  |  |
| CVS | **3.68%** | 3.08% | 1.50% | 2.27% | 2.67% | 1.85% |
| Infection | 0.25% | **0.77%** | 0.23% | 0.10% | 0.37% | 0.25% |
| Lung cancer | 80.15% | 85.38% | **90.76%** | 85.66% | 69.54% | 86.72% |
| Non-lung cancer | 2.45% | 0.77% | 1.53% | **2.27%** | 2.67% | 1.79% |
| Other cause | 13.48% | 10.00% | 5.98% | 9.69% | **24.76%** | 9.39% |
| Multinomial regression models |  |  |  |  |  |  |
| Missing COD | 29.78% | 26.15% | 10.06% | 17.51% | 34.84% | 15.36% |
| Lung Cancer | 69.24% | 72.31% | **89.12%** | 81.40% | 58.67% | 82.96% |
| Multiple COD | 0.98% | 1.54% | 0.83% | 1.09% | 6.49% | 1.68% |

Note: COD, causes of death.

**Figure 2.**
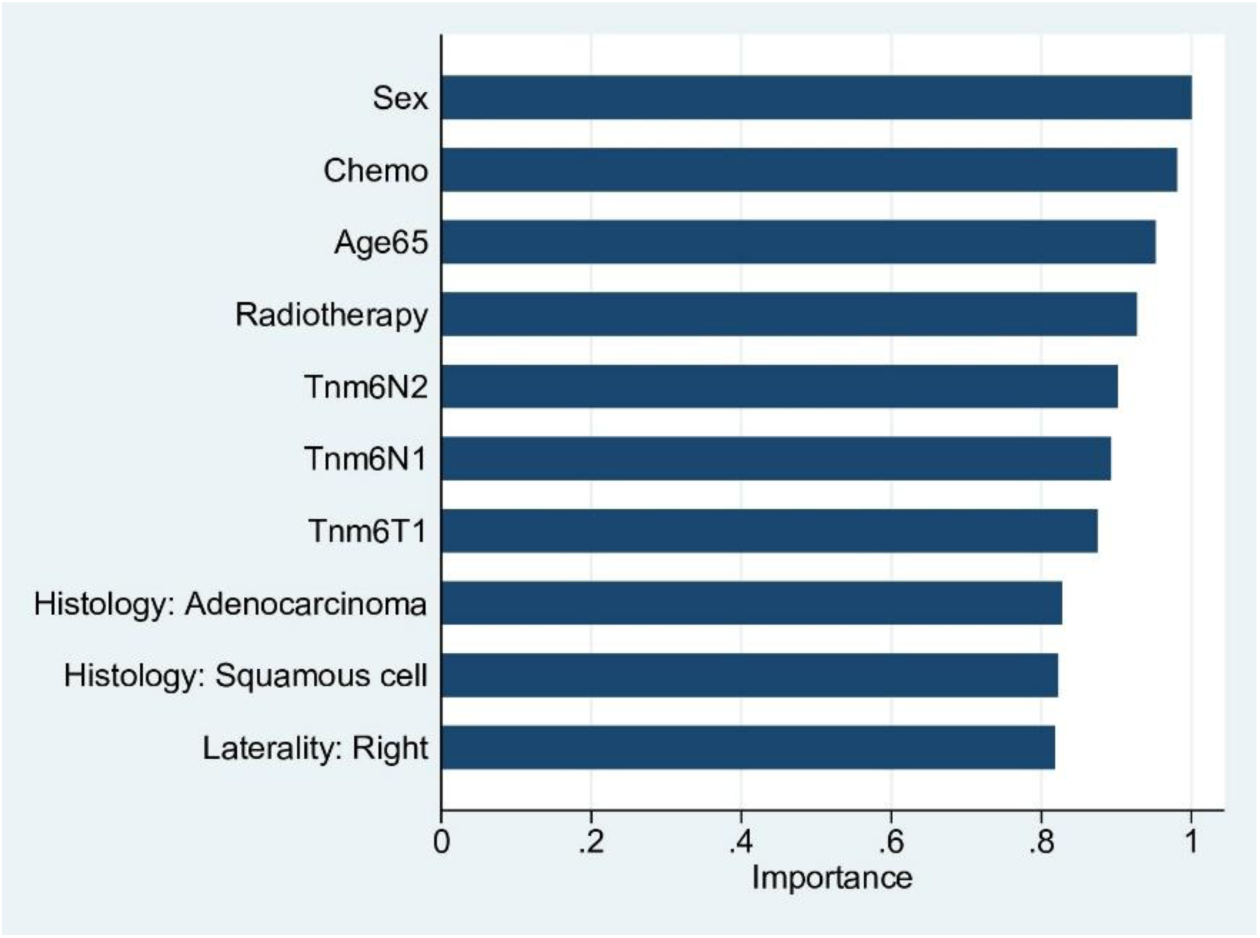
In the final random forest model of machine learning with 300 iteration, 10 variables and the highest accuracy (69.72%), the top-10 most important factors in the random-forest model were sex, chemotherapy status, age, radiotherapy status and nodal status. Confirmation by pathology was not in the selected 10 variables/factors.

Interestingly, the pathological confirmation of the diagnosis appeared not very importance in the model. In the multinomial regression model, we also found that sex, chemotherapy status, age (65+ vs <65 years), radiotherapy status, nodal status, T category, histology type and laterality (**Table 3**), while other factors were not, despite being associated with the multicategory COD in univariable regression analysis (Chi-squared test, Table 1). This finding was consistent with that from RF model.

**Table 3.** The factors associated with multicategory causes of death in lung cancer patients as shown in a multinomial regression model.

|  | CVS |  |  | Infection |  |  | Non_lung_cancer |  |  | Other cause |  |  |
| --- | --- | --- | --- | --- | --- | --- | --- | --- | --- | --- | --- | --- |
|  | OR | 95% CI | P | OR | 95% CI | P | OR | 95% CI | P | OR | 95% CI | P |
| **Male (vs female)** | 1.19 | (1.04 - 1.36) | 0.009 | 1.21 | (0.90 - 1.63) | 0.197 | 0.95 | (0.85 - 1.07) | 0.397 | 0.79 | (0.73 - 0.86) | <.001 |
| **Radiotherapy (vs none)** | 0.70 | (0.60 - 0.82) | <.001 | 0.47 | (0.32 - 0.71) | <.001 | 0.76 | (0.67 - 0.86) | <.001 | 0.53 | (0.47 - 0.59) | <.001 |
| **Chemotherapy (vs none)** | 0.61 | (0.51 - 0.71) | <.001 | 0.59 | (0.41 - 0.87) | 0.007 | 0.76 | (0.66 - 0.87) | <.001 | 0.60 | (0.54 - 0.67) | <.001 |
| **Age (65+ vs <65 yr)** | 1.83 | (1.54 - 2.17) | <.001 | 0.73 | (0.53 - 1.00) | 0.047 | 1.00 | (0.88 - 1.13) | 0.964 | 0.56 | (0.51 - 0.62) | <.001 |
| **Histology** |  |  |  |  |  |  |  |  |  |  |  |  |
| **Adenocarcinoma** | 0.73 | (0.61 - 0.86) | <.001 | 0.64 | (0.43 - 0.95) | 0.025 | 0.99 | (0.86 - 1.14) | 0.903 | 0.73 | (0.65 - 0.81) | <.001 |
| **Small cell carcinoma** | 0.63 | (0.48 - 0.84) | 0.001 | 0.84 | (0.47 - 1.49) | 0.554 | 0.67 | (0.54 - 0.83) | <.001 | 0.49 | (0.40 - 0.59) | <.001 |
| **Squamous cell carcinoma** | 1.12 | (0.95 - 1.34) | 0.183 | 0.95 | (0.64 - 1.41) | 0.797 | 0.82 | (0.70 - 0.98) | 0.026 | 0.75 | (0.67 - 0.85) | <.001 |
| **Other Histology** | - | - | - | - | - | - | - | - | - | - | - | - |
| **N category** |  |  |  |  |  |  |  |  |  |  |  |  |
| **N1 vs N0** | 3.18 | (1.65 - 6.12) | 0.001 | 7.51 | (1.38 - 40.90) | 0.02 | 1.43 | (0.87 - 2.34) | 0.158 | 2.08 | (1.40 - 3.09) | <.001 |
| **N2 vs N0** | 2.42 | (1.19 - 4.90) | 0.014 | 2.86 | (0.42 - 19.51) | 0.284 | 1.42 | (0.81 - 2.49) | 0.215 | 1.79 | (1.16 - 2.77) | 0.008 |
| **N3 vs N0** | 1.91 | (1.04 - 3.50) | 0.036 | 1.74 | (0.40 - 7.53) | 0.456 | 0.87 | (0.56 - 1.33) | 0.516 | 1.36 | (0.96 - 1.91) | 0.079 |
| **Percent of foreign-born residents, quartile** |  |  |  |  |  |  |  |  |  |  |  |  |
| **Q1 vs Q4** | 0.89 | (0.71 - 1.11) | 0.298 | 0.65 | (0.39 - 1.08) | 0.093 | 1.01 | (0.83 - 1.22) | 0.955 | 0.79 | (0.67 - 0.92) | 0.002 |
| **Q2 vs Q4** | 0.96 | (0.77 - 1.20) | 0.724 | 0.81 | (0.50 - 1.34) | 0.415 | 0.95 | (0.78 - 1.15) | 0.613 | 0.93 | (0.80 - 1.08) | 0.349 |
| **Q3 vs Q4** | 0.81 | (0.67 - 0.98) | 0.035 | 0.78 | (0.51 - 1.19) | 0.242 | 0.84 | (0.71 - 0.99) | 0.037 | 0.91 | (0.80 - 1.03) | 0.14 |
| **Laterality** |  |  |  |  |  |  |  |  |  |  |  |  |
| **Bilateral (vs Right side)** | 0.66 | (0.32 - 1.36) | 0.261 | 2.49 | (1.06 - 5.88) | 0.037 | 1.62 | (1.14 - 2.31) | 0.007 | 0.83 | (0.53 - 1.28) | 0.397 |
| **Left side (vs Right side)** | 1.06 | (0.93 - 1.21) | 0.404 | 0.84 | (0.63 - 1.14) | 0.272 | 0.96 | (0.86 - 1.08) | 0.504 | 0.99 | (0.91 - 1.09) | 0.884 |

Note: OR, odds ratio; CI, confidence interval.

## Discussion

In this population-based study of 40,000 lung cancer patients with more than 12 years of follow-up, we optimized a RF model of machine learning to predict the specific cause of death in lung cancer patients. The RF model also appears more accurate than multinomial regression model (69.8% vs 64.6%, P<0.001). We also identified the factors linked to the 5-category COD in lung cancer patients, which was not reported by previous works [4, 7].

This study is limited by using the cases of a single diagnosis year, while we feel the large number of cases justified for our findings. Future studies may include the cases of recent diagnosis years so that the findings will be more related to the recent cases. Moreover, the dataset appeared unbalanced in the outcome. Such a nature of the dataset is the real-world evidence, but posed a challenge to modelling. This in part explained why the multinomial regression model had many cases missing an assigned COD. Finally, the findings were not validated by an external dataset. Future works should be carried out using other dataset to validate our findings.

In summary, we here identified the factors that were independently associated with 5-category long-term COD in lung cancer patients in the USA. We also tuned the RF model and showed that it was significantly more accurate in predicting 5-category long-term COD.

## Data Availability

The SEER data were available upon request to the SEER website (www.seer.cancer.gov). All other data are available upon request.

